# BA.4/BA.5 early surge report Austria

**DOI:** 10.1101/2022.07.12.22277534

**Authors:** Alexander Gamisch, Maria Elisabeth Mustafa

## Abstract

More than two years into the COVID-19 pandemic, the emergence of the Omicron subvariants BA.4 and BA.5 initiated the fifth wave. BA.4/BA.5 share identical spike proteins and are thus difficult to differentiate using standard diagnostic tests. The true frequency and diversity of the two variants in Austria is therefore largely unknown. Aim of this report is to take stock about the frequency and diversity of BA.4 & BA.5 and their subvariants based on whole genome sequencing (WGS) data as deposited at GISAID database. Results show that most sequenced cases belong to BA.5 (c. 86 %) rather than BA.4 (c. 14 %) and that most of the global diversity (24 out of 32 subvariants) of BA.4 and BA.5 in terms of subvariants described is already present in Austria. However, most cases can be attributed to a few subvariants (e.g. BA.5.1, BE.1.1, BA.5.3, BA.5.2.1) with high estimated growth advantage over BA.2 (ranging 103 to 159 %) and may be worth monitoring as the immediate wave unfolds.

## Introduction

At the dawn of 2022, more than two years into the COVID-19 pandemic, the emergence of the Omicron subvariants BA.4 and BA.5 in South Africa initiated the fifth wave (Tegally et al. 2022, Mohapatra et al. 2022). In Austria BA.4/BA.5 was first detected mid-May (calendar week 15) and became dominant within weeks in combination with rising cases numbers (AGES^1^) thereby dampen any hope for a “summer break” before autumn. BA.4/BA.5 carry additional spike mutations (69-70del, L452R, F486V and the wild type amino acid at Q493) than BA.2 and are likely more transmissible than the latter due to immune evasion (Tegally et al. 2022; Hoteit and Yassine 2022; Hachmann et al. 2022). BA.4/BA.5 are further hardly distinguishable by PCR based screening tests and partial genome sequencing approaches that commonly target the spike gene region (cf. O’Toole et al. 2022; ECDC 2022). As a corollary, the true frequency and diversity of the two variants in Austria is largely unknown. Moreover, at the time of writing 32 more obscure subvariants of BA.4 and BA.5 have already been described^2^. These subvariants usually represent epidemiological significant clusters with a significant amount of onward transmission that are defined by at least one unambiguous evolutionary event (viz. mutation; Rambaut et al. 2020). Most mutations are inconsequential, but some do alter the biological properties of the virus (e.g. Saito et al. 2022; Tegally et al. 2022; Hoteit and Yassine 2022; Hachmann et al. 2022). Therefore, it is useful to track the frequency of the mutations/subvariants when they emerge. Aim of this report is to provide an overview about the frequency and diversity of BA.4 & BA.5 and their subvariants in Austria based on public available whole genome sequencing (WGS) data as deposited at GISAID database (Elbe & Buckland-Merrett 2017). The potential risk of these subvariants in terms of relative growth advantage versus a BA.2 baseline is assessed.

## Methods

A total of 1199 complete genomes of BA.4 and BA.5 collected in Austria have been downloaded from GISAID (Elbe & Buckland-Merrett 2017; GISAID accessed on 08.07.22). Sequences have then been classified to the Pango-nomenclatur system (Rambaut et al. 2020) using the Nextclade online tool^3^ v.2.0.0 (Aksamentov et al. 2021) and the frequencies per variant over sampling time were recorded. The Nextclade mutation profile was further used to manually designate novel variants^4^ not yet recognized by the software. Finally, the Covspectrum^5^ (Chen et al. 2022) platform was used to query GISAID genomic data for global variant frequencies and to calculated transmission advantage versus a BA.2 baseline following Chen et al. (2021). Due to the limited number of Austrian BA.4/BA.5 genomes (n =1199; see also Tab.1) the relative growth advantage per week (in percentage; 0% means equal growth) in the timeframe of the last 3 months [2022-04-04 to 2022-07-04] was calculated based on European genomes.

**Table 1.** BA.4 and BA.5 subvariants, frequency, associated mutations, genome frequency global and in Austria as well as growth advantage ersus BA.2 based on genomic sequencing data.

| Subvariant | Initial predominant countries<sup>a</sup> | Associated mutation(s)<sup>a</sup> | Global<sup>b</sup> | Austria | Current adv. per week (%)<sup>b</sup> | Conf. int. (%)<sup>b</sup> |
| --- | --- | --- | --- | --- | --- | --- |
| BA.4 | South Africa | parental lineage | 26452* | 164* | 93% | 91-95% |
| BA.4.1 | South Africa | T21570G (S:V3G) | 16194* | 41 | 84% | 82-87% |
| BA.4.1.1 | South Africa and Germany | A23570G (S:I670V) | 938 | 18 | 76% | 69-83% |
| BA.4.1.2 | Austria | C3176T (Orf1a:P971S) | 168 | 41 | 21% | 14-27% |
| BA.4.1.3 | Israel | A2435G (Orf1a:R724G) | 122 | 0 | NA | NA |
| BA.4.1.4 | Israel | G25266T (S:C1235F) | 320 | 0 | NA | NA |
| BA.4.2 | USA | C5173A (Orf1a:H1636Q) | 767 | 1 | 107% | 84-130% |
| BA.4.3 | Israel | C21365T (Orf1b:P2633L) | 169 | 0 | 72% | 37-106% |
| BA.4.4 | USA | C25708T (Orf3a:L106F) | 1491 | 7 | 101% | 92-111% |
| BA.4.5 | Australia | G22104A (S:G181E) | 154 | 0 | NA | NA |
| BA.5 | South Africa | parental lineage | 65951* | 1035* | 112% | 111-114% |
| BA.5.1 | Portugal | C29666T (ORF10:L37F) | 23037* | 357 | 103% | 102-105% |
| BA.5.1.1 | USA | C25672A (Orf3a:L94I) | 806 | 0 | NA | NA |
| BA.5.1.2 | Europe | G27214A (Orf6:V5I) | 270 | 3 | 151% | 131-172% |
| BA.5.1.3 | Germany Portugal and Spain | G22427A (S:V289I) | 709 | 13 | 114% | 105-122% |
| BA.5.2 | South Africa, England and USA | A28330G (ORF9b:D16G) | 21221* | 78 | 133% | 130-136% |
| BA.5.2.1 | South Africa, England and USA | A27038G (ORF5:T172T) | 12918 | 106 | 125% | 122-128% |
| BA.5.2.2 | France, Martinique | G20356A (Orf1b:G2297S) | 257 | 0 | 85% | 74-96% |
| BA.5.2.3 | UK | G625T (Orf1a:K120N) | 334 | 2 | 175% | 154-196% |
| BF.1 | England, Denmark, Spain and Scotland | G21520T (Orf1ab:V7086F)<br>(Orf1b:2685F) | 1944* | 0 | 90% | 85-94% |
| BF.1.1 | USA | G10835A (Orf1a:V3524I) | 191 | 29 | 75% | 41-109% |
| BF.2 | Denmark | G14398T (Orf1b:V311L) | 260 | 2 | 122% | 106-137% |
| BF.3 | India | C23202T (S:T547I) | 137 | 0 | NA | NA |
| BF.4 | England | A22337G (S:T259A) | 248 | 0 | 134% | 115-153% |
| BA.5.3 | Germany and South Africa | C1931A (ORF1ab:Q556K) | 10338* | 112 | 105% | 103-108% |
| BA.5.3.1 | South Africa, Austria and England | G28681T (N:E136D) | 1933 | 6 | 150% | 145-154% |
| BA.5.3.2 | Germany | G621A (ORF1ab:R119H) | 784 | 11 | 67% | 62-71% |
| BA.5.3.3 | UK | C6843T (Orf1a:S2193F) | 369 | 2 | 118% | 105-130% |
| BA.5.3.4 | Germany | G20718T (Orf1b:M2417I) | 267 | 1 | 64% | 56-71% |
| BE.1 | South Africa, Austria and England | G16935A (ORF1b:M1156I) | 7321* | 55 | 154% | 149-159% |
| BE.1.1 | Germany | C11750T (Orf1a:L3829F) | 3817 | 163 | 159% | 153-165% |
| BE.2 | Belgium | C2910T (Orf1a:T882I) | 110 | 0 | 66% | 53-80% |
| BA.5.5 | USA | C21789T (S:T76I) | 6417 | 12 | 142% | 127-157% |
| BA.5.6 | USA | C17402T (Orf1b:A1312V) | 1831 | 5 | 169% | 144-194% |

<sup>a</sup>https://www.pango.network/summary-of-designated-omicron-lineages/ [accessed 04.07.22]
<sup>b</sup>https://cov-spectrum.org [accessed 04.07.22]
\*including subvariants

## Results and Discussion

### BA.4 versus BA.5

All BA.4 and BA.5 subvariants exhibit strong relative growth advantage versus BA.2 (ranging between 21% and 175%; see Tab. 1) on a continental scale. For comparison, the Alpha variant (B.1.1.7) exhibit a growth advantage of 43–52 % compared to the “Wild type” SARS-CoV 2 strain (Chen et al. 2021). However, growth advantage of BA.4 subvariants is tendentially lower than those of BA.5 subvariants (Tab.1). Consequently, albeit BA.4 initially was more frequent in Austria than BA.5 the proportion has shifted in favor of BA.5 in recent weeks (around calendar week 17) (Fig.1). Now, in concordance with the global trend, the BA.5 variant is dominant with c. 86 % (1035/1199) compared to BA.4 (c. 14 % (164/1199) in Austria (Fig.1).

**Figure 1.**
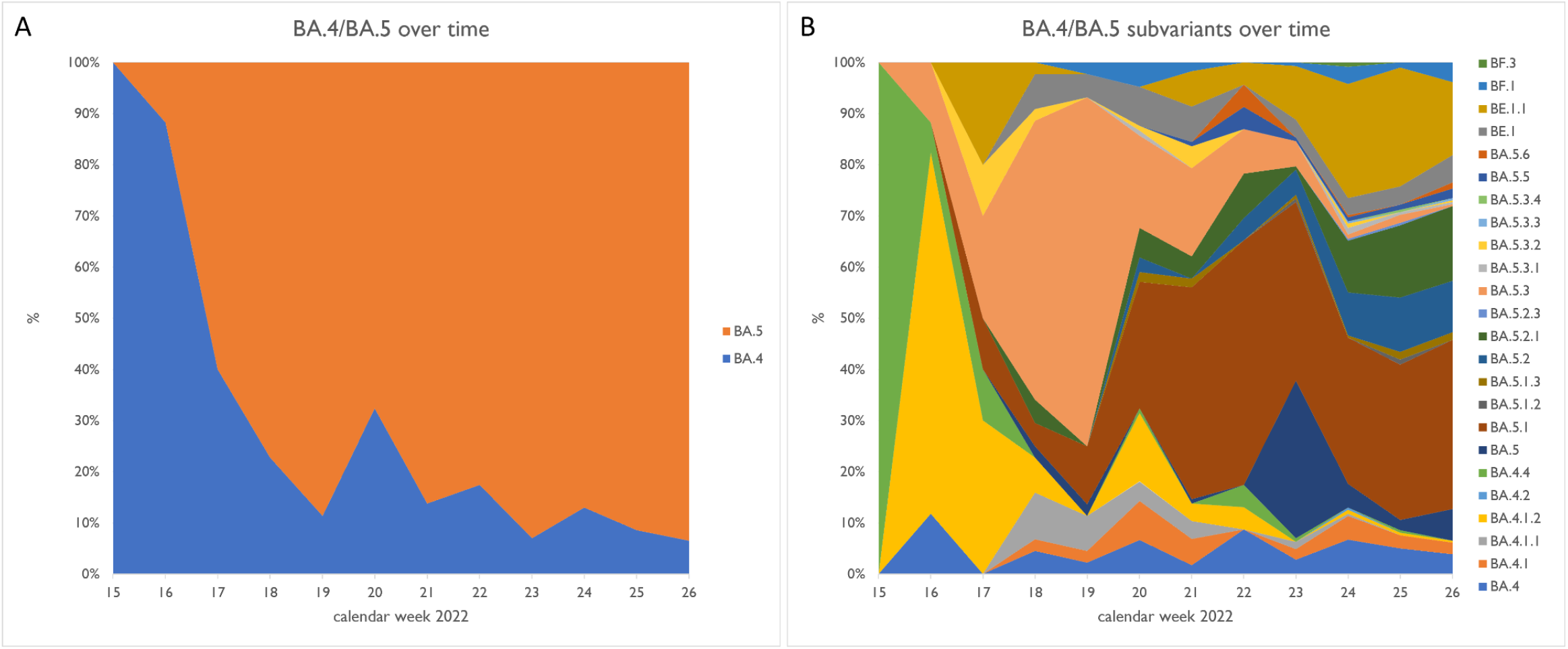
Omicron subvariants BA.4 and BA.5 genomes over time (calendar weeks) in Austria. A, BA.4 and BA.5 including subvariants over me. B, subvariants of BA.4 and BA.5 over time.

### BA.4/BA5 subvariants

From the 32 subvariants of BA.4/BA5 a total of 24 subvariants were already detected in Austria over the past 12 weeks (calendar week 15 to 26) (Fig.1, Fig.2) likely due to multiple independent introductions of the different strains. In Austria, the top five most frequently sequenced subvariants over the whole timeframe are BA.5.1, BE.1.1, BA.5.3, BA.5.2.1 and BA.5 thereby representing a total of c. 68 % of cumulative cases sequenced. Of those BA.5.1, BE.1.1 and BA.5.2.1 show signs of sustained growth whereas BA.5.3 apparently peaked in calendar week 19 and is on decline ever since. The top five subvariants with the highest growth advantage (ranging between 175 and 151 %) are BA.5.1.2, BE.1, BE.1.1, BA.5.6, BA.5.2.3 whereas three of those (BA.5.6, BA.5.2.3, BA.5.1.2) are only known from a handful of recent cases in Austria so far (Tab.1, Fig. 3). The variants BE.1.1, BA.5.2.1, BA.5.1 on the other hand appear to be more successful than their peers when judged by cases numbers and estimated growth advantages (Tab.1, Fig. 3) and may be worth monitoring as the proximate wave unfolds.

**Figure 2.**
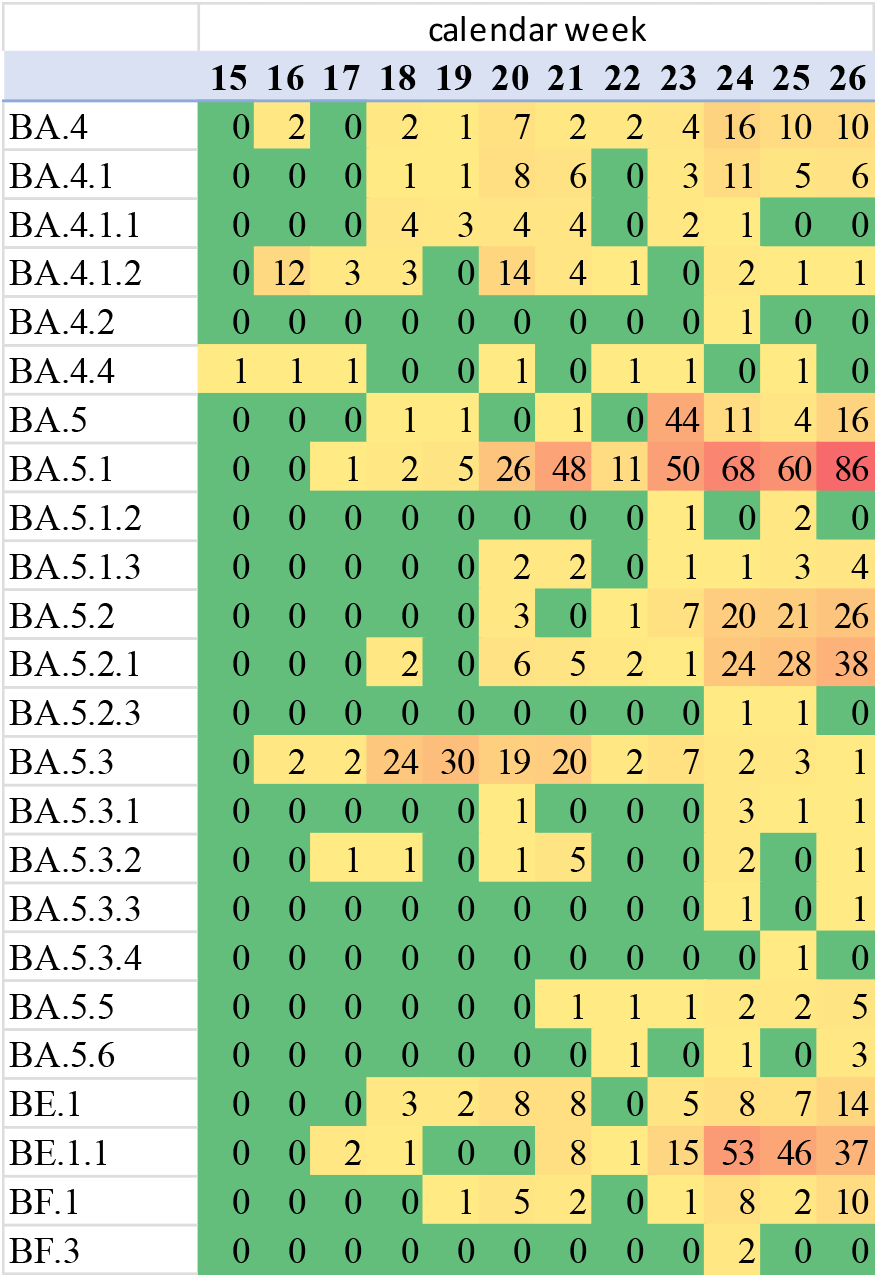
Heatmap of number BA.4 and BA.5 genomes over time (between calendar week 15 and 26) in Austria.

**Figure 3.**
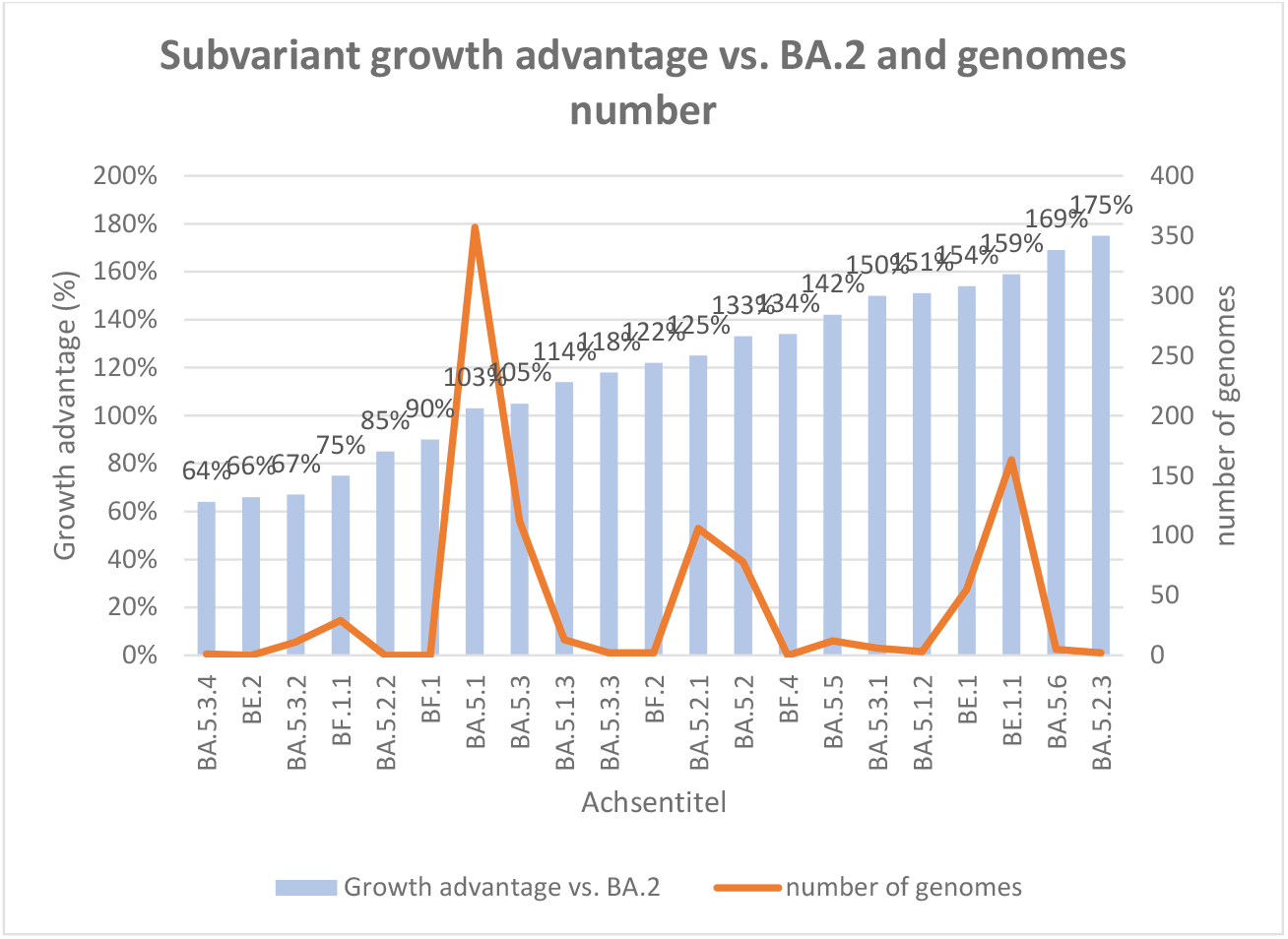
Subvariant growth advantage vs. BA.2 and genomes number.

## Data Availability

All data produced in the present study are available upon reasonable request to the authors

## Acknowlegements

We gratefully acknowledge all data contributors, i.e. the authors and their originating laboratories responsible for obtaining the specimens, and their submitting laboratories for generating the genetic sequence and metadata and sharing via GISAID. Following labs submitted to GISAID: Bergthaler laboratory, CeMM Research Center for Molecular Medicine of the Austrian Academy of Sciences; Lifebrain Covid Labor GmbH; Dr. Mustafa, Dr. Richter Labor für medizinisch-chemische und mikrobiologische Diagnostik GmbH, Abteilung Molekularbiologie; Center for Virology, Medical University of Vienna; Institute of Virology Department of Hygiene, Microbiology and Public Health at Innsbruck Medical University.

## Footnotes

1 https://www.ages.at/mensch/krankheit/krankheitserreger-von-a-bis-z/coronavirus#c12422 [accessed 04.07.22]

2 https://github.com/cov-lineages/pango-designation [accessed 04.07.22]

3 https://clades.nextstrain.org [accessed 08.07.22]

4 https://www.pango.network/summary-of-designated-omicron-lineages/ [accessed 04.07.22]

5 https://cov-spectrum.org [accessed 04.07.22]

